# Informal Payments in Ukrainian Primary Healthcare System: cross-sectional study, 2022-2023 years

**DOI:** 10.1101/2025.09.22.25336385

**Authors:** Maksym Duda, Nataliia Kovalenko, Anna Tokar, Oleh Sorotsynskyi, Paola Pavlenko, Vladyslav Odrynskyi

## Abstract

Countries with emerging and developing economies, particularly during crises like the ongoing full-scale war in Ukraine, face significant challenges in achieving universal health coverage. This secondary data analysis focused on informal payments (IPs) in Ukraine’s public primary healthcare (PHC) system, using a dataset from a study that examined IPs across four priority healthcare services: childbirth, neonatal care, myocardial infarction, and stroke care. Conducted from June 1, 2022 till May 31, 2023, across all Ukrainian regions (excluding temporarily occupied territories), the study analysed anonymous surveys of 2,031 patients using multivariable regression in SPSS. We identified that women and individuals from larger households were more likely to make IPs at the PHC level. Older patients generally viewed IPs negatively, though age did not significantly correlate with IP behaviour. Lower-income individuals also viewed IPs negatively. Resistance to health system reforms was associated with more positive attitudes towards IPs, with regional variations observed. The study results will support the Government’s evidence-based policy decisions and data-driven interventions that reduce corruption and improve health outcomes in Ukraine.

## Introduction

Informal payments (IPs) for healthcare services are a well-documented phenomenon in countries with emerging and developing economies [1–6], including those in Eastern and Central Europe and Central Asia. Thus, a range of empirical studies have examined IPs, focusing on their trends, patterns, policy implications, motivations, and associated attitudes and perceptions [3, 4, 7–16]. Overall, IPs pose challenges for patients, healthcare providers, and the health in general system [1, 2, 5–15, 17, 18], particularly during crisis periods such as the COVID-19 pandemic [15], military conflicts, wars and post-wars periods [3, 19–21].

Prior to the full-scale war in Ukraine, paying for healthcare services through IPs was common, with a number of studies demonstrating their high prevalence and overall acceptance by both healthcare providers and patients [8, 11, 16, 22–25]. According to Ukraine’s Constitution [26], citizens are entitled to healthcare free of charge in public facilities. However, inherited from the Soviet Union Semashko system demonstrated underfunding, overblown and redundant network of facilities, non-motivational financing principles, lack of financial risk protection, which has resulted in public facilities building up revenue by charging IPs to patients.

In response, Ukraine has embarked on significant health system reforms, as outlined in Concept on Health Care Financing Reform [27] and in the Law on State Financial Guarantees of Health Care Services to the Population [28], approved by the Parliament in 2016-2017 and effective from 2018. The reform addressed, among several core problems, the high level of out-of-pocket payments (OOPs) as the share of health spending in Ukraine, reaching 49% of total expenditure (2018), a figure that places the country among those in Europe with the highest levels of OOPs as part of healthcare expenditure[29, 30] .

These reforms aim to transition Ukraine towards real universal health coverage (UHC), compared to formally declared before, as well as improve efficiency and equity in public spending. Key focus is made on the creation of a single national purchaser (National Health Service of Ukraine, NHSU), more transparent resource allocation, strategic purchasing with a purchaser–provider split, explicitly defined health benefits, and new payment methods for healthcare providers [29, 31, 32]. Annually NHSU manages a state-funded Program of Medical Guarantees with more than 4 billion USD (2024) budget. It is expected that these reforms will, over time, reduce the prevalence of IPs and improve access to healthcare services promoting equity and UHC.

Since 2020, the healthcare system of Ukraine has faced significant challenges, including the COVID-19 pandemic and the full-scale war in Ukraine. They had a substantial impact on the healthcare system, complicating the roll-out and scaling of the reforms and exacerbating existing healthcare disparities.

This study was designed and formulated to investigate informal payments (IPs) within Ukraine’s public primary healthcare system, with a focus on factors associated with lifetime occurrences of such payments, recent IPs within the past 12 months, and prevailing attitudes toward this practice.

## Materials and Methods

### Study setting and participants

We performed secondary analysis of cross-sectional data collected within the study *“Informal Payments for Four Priority Services under the Program of Medical Guarantees at the Specialized Healthcare Level”*. The study considered question regarding IPs at specialized healthcare level with additional topic regarding primary healthcare (PHC), expectation, and perceptions. The initial study and analyses were performed by the Kyiv School of Economics Institute, and USAID Health Reform Support, Kyiv Ukraine [33].

In this paper, we specifically focus at the PHC level, which serves in most cases as the critical first point of contact for patients before they access specialized care. This aspect was not addressed in the primary data analysis, which centered on the four priority healthcare services.

The study methodology was informed by findings of a systematic reviews focusing on research methods and instruments used to examine IPs for healthcare services [34, 35].

The range of forms that IPs can take presents a challenge to providing a comprehensive definition. Nonetheless, we were guided by a systematic review on defining IPs in healthcare [17] and, thus, in this study, IPs are defined as follows:

1. Monetary payments or equivalents given to doctors or other healthcare facility personnel by patients or their relatives.
2. Donations solicited or demanded from patients or their relatives.
3. Purchases of medicines and/or medical devices based on a list provided by the healthcare facility that were to be covered by the state funding under the Program of Medical Guarantees
4. In-kind gifts such as goods or services offered to healthcare providers by patients, or their relatives.
5. Any payments not specified in official healthcare facility regulations.

The field phase covered the period from June 1, 2022, to May 31, 2023. The study proceeded with data validation, rigorous analysis, and comprehensive reporting.

The study was conducted across all regions of Ukraine, excluding temporarily occupied territories. The sample size was calculated based on data from the ’Center for Medical Statistics of the Ministry of Health of Ukraine,’ specifically concerning the four priority services over the past 12 months: childbirth, neonatal care, acute myocardial infarction, and acute stroke care. Survey sites were selected to include geographically representative sample of patients.

Data were collected across five macro-regions in Ukraine, following the regional distribution criteria established by the NHSU [36], in particular: North (Poltavska, Sumska, Kharkivska and Chernihivska oblasts), South (Mykolayivska, Odeska, Khersonska oblasts, and Autonomous Republic of Crimea), West (Volynska, Ivano-Frankivska, Zakarpatska, Lvivska, Rivnenska, Ternopilska, Khmelnytska, Chernivetska oblasts), East (Dnipropetrovska, Donetska, Zaporizhska, Kirovohradska, Luhanska oblasts) and Center (Kyiv city and Kyivska, Vinnytska, Zhytomyrskа, Cherkaska oblasts).

Eligibility criteria for participants included being 18 years of age or older, holding Ukrainian citizenship, providing both written and verbal consent, and receiving at least one of the four priority healthcare services (childbirth, neonatal care, acute myocardial infarction, or acute stroke care) at a public healthcare facility during June 1, 2022-May 31, 2023.

Overall, 2,031 respondents participated in the survey. The selection of priority healthcare services, particularly childbirth and neonatal care, influenced the gender distribution in the study structure, leading to a higher number of female participants (77.4%, N= 1,031). Additionally, Ukraine has historically maintained a high percentage of university-educated individuals [17], closely aligning with the sample composition in our study (59.2%, N=1,209). Details of socio-demographic variables are provided in (Table 1).

**Table 1.** Descriptive characteristics of variables.

| Variable | Value (N) | Frequency (%) |
| --- | --- | --- |
| **Outcomes** |  |  |
| **Ever in life IPs** |  |  |
| **Have you made any IPs for services provided by family doctors/primary care ever in life?** |  |  |
| No |  |  |
| Yes | 1041 | 51.3% |
| Missing | 990 | 48.7% |
|  | 0 | 0% |
| **IPs within last 12 months** |  |  |
| **Have you made any IPs for services provided by family doctors/primary care between June 1, 2022, and May 31, 2023?** |  |  |
| **year** |  |  |
| No |  |  |
| Yes | 1041 | 51.3% |
| Missing | 925 | 45.5% |
|  | 65 | 3.2% |
| **Patient Attitudes Toward IPs** |  |  |
| Negative | 1270 | 62.5% |
| Positive and neutral | 727 | 35.8% |
| Missing | 34 | 1.7% |
| **Explanatory variables** |  |  |
| **Sex** |  |  |
| Man | 458 | 22.6% |
| Female | 1573 | 77.4% |
| Missing | 0 | 0% |
| **Age in years** |  |  |
| 18-39 years old | 1096 | 54% |
| 40-64 years old | 681 | 33.5% |
| 65+ years old | 254 | 12.5% |
| Missing | 0 | 0% |
| Median (IQR) | 37 (30-57) |  |
| **Belonging to a vulnerable group** |  |  |
| Yes | 1478 | 72.8% |
| No | 553 | 27.2% |
| Missing | 0 | 0% |
| **Education level** |  |  |
| ≤ 11 years of schooling | 268 | 13.3% |
| Technical College or Vocational school | 554 | 27.3% |
| University degree and postgraduate degree | 1205 | 59.2% |
| Missing | 4 | 0.2% |
| **Number of People in Household** |  |  |
| 1-2 people | 611 | 30.1% |
| 3-4 people | 1199 | 59% |
| 5+ people | 221 | 10.9% |
| Missing | 0 | 0% |
| **Income Level of the Household** |  |  |
| Can afford only the most necessary products | 773 | 38.1% |
| Can afford small additional items | 857 | 42.2% |
| Can afford everything | 401 | 19.7% |
| Missing | 0 | 0% |
| **Employment status: Full-time work** |  |  |
| No | 1283 | 63.2% |
| Yes | 748 | 36.8% |
| Missing | 0 | 0% |
| **Employment status: Part-time work** |  |  |
| No | 1908 | 93.9% |
| Yes | 123 | 6.1% |
| Missing | 0 | 0% |
| **Employment status: Self-employed** |  |  |
| No | 1953 | 96.2% |
| Yes | 78 | 3.8% |
| Missing | 0 | 0% |
| **Employment status: Pensioner** |  |  |
| No | 1700 | 83.7% |
| Yes | 331 | 16.3% |
| Missing | 0 | 0% |
| **Settlement size** |  |  |
| Regional center | 1392 | 68.5% |
| City with more than 100 thousand inhabitants | 325 | 16% |
| Settlement with less than 100 thousand inhabitants | 314 | 15.5% |
| Missing | 0 | 0% |
| **Macro-region** |  |  |
| North | 336 | 16.5% |
| South | 239 | 11.8% |
| West | 517 | 25.5% |
| East | 476 | 23.4% |
| Center | 461 | 22.7% |
| Missing | 2 | 0.1% |
| **Changes of Residency due to the Full-scale war** |  |  |
| Yes | 261 | 12.8% |
| No | 1766 | 87.0% |
| Missing | 4 | 0.2% |
| **Changes in Access to Healthcare after Full-Scale War (24.02.2022)** |  |  |
| Yes, definitely in positive way | 216 | 10.6% |
| Yes, probably in positive way | 632 | 31.2% |
| No changes | 809 | 39.8% |
| No, probably in negative way | 145 | 7.1% |
| No, definitely in negative way | 79 | 3.9% |
| Difficult to say | 150 | 7.4% |
| Missing | 0 | 0% |
| **Changes in IPs in Healthcare after Full-Scale War (24.02.2022)** |  |  |
| Yes, definitely in positive way | 201 | 9.9% |
| Yes, probably in positive way | 601 | 29.6% |
| No changes | 774 | 38.1% |
| No, probably in negative way | 110 | 5.4% |
| No, definitely in negative way | 109 | 5.4% |
| Difficult to say | 236 | 11.6% |
| Missing | 0 | 0% |
|  | 0 | 0% |
| **Perceived success of healthcare reform in eliminating IPs** |  |  |
| Definitely possible | 205 | 10.1% |
| Probably possible | 747 | 36.8% |
| Maybe possible, maybe no | 502 | 24.7% |
| Probably impossible | 212 | 10.4% |
| Definitely impossible | 249 | 12.3% |
| Difficult to say | 116 | 5.7% |
| Missing | 0 | 0% |

### Study design

Interviewers with prior survey experience were recruited and trained in the study’s methodologies and tools.

Participants completed an interviewer-administered anonymous standardized questionnaire that collected information on Section 1: Basic information about the respondent; Section 2: Perception of informal payments for healthcare services in general; Section 3: Informal payments in selected healthcare facilities; Section 3a: Maternity care; Section 3b: Medical care for new-borns in complicated neonatal cases; Section 3c: Medical care for acute myocardial infarction; Section 3d: Medical care for acute stroke; and Section 4: Patient satisfaction (Appendix A). The questionnaire underwent pre-testing to determine the optimal number of questions and to refine question phrasing, aiming for an interview duration of approximately 45 minutes.

### Ethics

Participation in the study was entirely voluntary, with all participants providing both written and verbal consent. Approximately 30% of eligible participants chose not to participate, primarily due to psychological barriers in discussing informal payments and limited knowledge of medical guarantee programs. Some respondents were unreachable due to health conditions or opted out to avoid revisiting traumatic experiences. In some cases, relatives of patients were interviewed as substitutes, with the patients’ permission.

Ethical approval was secured from the Kyiv School of Economics (Appendix B).

### Outcome variables

We examined IPs using three binary outcome variables (0 = No, 1 = Yes):

1. Self-reported IPs provided at any point in life for services received from family doctors or at the PHC level.
2. Self-reported IPs during the period from June 1, 2022, to May 31, 2023 (in the last 12 months), for services received from family doctors or at the PHC level.
3. A positive attitude towards IPs.

### Explanatory variables

The selection of explanatory variables was constrained by the available data. Our choices were guided by a scoping literature review conducted by our team. Consequently, we examined the following variables: age, gender, education, employment status, status as a vulnerable group, income level, number of people living in the household, and region of residence, size of the settlement, changed residency due to war, perceived changes in IPs in healthcare after full-scale war and changes in access to healthcare after full-scale war, perceived success of healthcare reform in eliminating IPs.

### Statistical analysis

We calculated descriptive statistics and excluded responses classified as ‘hard to answer,’ ‘I do not know/remember,’ or ‘refuse to answer’ as missing data. Such missing data were not included in the bivariate or multivariable analyses, and the presence of missing data is noted in the tables.

In the bivariate analyses, we compared categorical variables using the chi-square test. Multivariable logistic regression was employed to examine the associations between explanatory variables and the three outcome variables: lifetime occurrences of informal payments on PHC level, recent IPs within the past 12 months on PHC level, and prevailing attitudes towards this practice.

For the multivariable logistic regression analysis, we utilized a backward stepwise selection technique with the Wald chi-square test to determine the final model of best fit. Explanatory variables were removed one at a time if they did not show an association with the outcome at a 5% significance level.

All analyses were conducted using SPSS (version 26). We present all bivariate and multivariable results in the Results section (Table 1).

### Limitations

Approximately 30% of eligible participants declined to participate in the surveys. These individuals may differ significantly from those who chose to participate.

Another limitation of the study is the potential for informational and recall biases inherent in self-reported data, especially regarding sensitive questions about IPs. Nevertheless, we contend that these biases were likely minimized through the use of trained interviewers, who helped ensure the accuracy and reliability of the responses.

Additionally, the cross-sectional nature of our study prevents us from establishing causal relationships between the outcomes and explanatory variables.

## Results

### Descriptive characteristics

Sociodemographic characteristics are shown in Table 1. Overall, the median age of participants was 37 years (IQR: 30 – 57), with more than three-quarters of them being females (77.4%) and belonging to at least one vulnerable group (72.8%). The majority of participants held a university or postgraduate degree (59.2%). Specifically, only 13.3% did not pursue higher education, while 27.3% graduated from a technical college or vocational school. Most participants’ households (59%) comprised 3–4 people, while 30.1% lived in smaller households with 1–2 people, and only a small proportion (10.9%) resided in households with 5 or more members. About 38.1% of participants could afford only the most essential products, while 42.2% could purchase small additional items. In contrast, one in five participants (19.7%) reported being able to afford everything, or almost everything. One-third (36.8%) were employed full-time, 6.1% worked part-time, 16.3% were retired and not working, and 3.8% were self-employed.

Most participants resided in oblast centers, including capital Kyiv city (68.5%); while roughly equal proportions lived in cities with populations over 100,000 (16%) and in smaller settlements (15.5%). Near one-fourth of participants lived in the western (25.5%), eastern (23.4%), and central (22.7%) parts of Ukraine, with fewer living in the northern (16.5%) and southern (11.8%) macro-regions. Around 13% were forced to change their residency due to the full-scale war.

Approximately 11% of participants reported negative changes in healthcare access after the onset of the full-scale war (7.1% probably negative, 3.9% definitely negative). Around 40% observed no changes, while a similar proportion reported positive changes (31.2% probably positive, 10.6% definitely positive). Regarding IPs, 38.1% of participants noted no changes since the full-scale war began. Positive changes were reported by 9.9% (definitely positive) and 29.6% (probably positive), while 5.4% in each group noted probably negative or definitely negative changes.

About one-quarter of participants (24.7%) were uncertain about the possibility of eliminating IPs in healthcare in Ukraine. Most believed it would be possible (10.1% definitely, 36.8% probably), yet a smaller proportion thought it would be probably impossible (10.4%) or definitely impossible (12.3%).

### Informal payment ever in life: Have you made any informal payments for services provided by family doctors/primary care ever in life?

48.7% of participants (95% CI: 46.5–50.9) reported having made IPs for at least once at some point in their lives.

In the bivariate model (Table 2), the following factors were associated with ever in life IP on PHC level: female participants (OR 1.3, 95% CI 1.1-1.6); households with 3-4 people (OR 1.4, 95% CI 1.1-1.7); lived in southern (OR 1.6, 95% CI 1.1-2.1), western (OR 2.0, 95% CI 1.6-2.6), eastern (OR 2.0, 95% CI 1.6-2.6) macro-regions of Ukraine; experienced changes in access to healthcare after full-scale war started in probably negative way (OR 1.9, 95% CI 1.2-2.8), in definitely negative way (OR 2.1, 95% CI 1.2-3.5), faced no changes (OR 1.6, 95% CI 1.2-2.1); faced changes in IPs since the full-scale war started in probably positive way (OR 1.5, 95% CI 1.1-2.1), probably negative way (OR 3.2, 95% CI 1.7-5.3), in definitely negative way (OR 3.4, 95% CI 2.1-5.6), no changes (OR 2.4, 95% CI 1.7-3.3), difficult to say (OR 1.7, 95% CI 1.1-2.4); evaluate expected overcoming of IPs in healthcare as probably possible (OR 1.5, 95% CI 1.1-2.0), probably impossible (OR 2.6, 95% CI 1.7-3.8), definitely impossible (OR 3.7, 95% CI 2.5-5.4), maybe possible, maybe no (OR 2.1, 95% CI 1.5-2.9), difficult to say (OR 1.7, 95% CI 1.1-2.7).

**Table 2.** Bivariate and Multivariable Analysis of Ever in Life Informal Payments for Primary Health Care Services Among Patients in Ukraine.

| Independent variables |  | Bivariate analysis |  |  | Multivariable analysis |  |  |
| --- | --- | --- | --- | --- | --- | --- | --- |
|  |  | N (%) | OR (95% CIs) | p-value | N (%) | AOR (95% CIs) | p-value |
| Age in years | 18-39 | 1096 (54%) | 1.0 (ref) |  | 1096 (54%) | 1.0 (ref) |  |
|  | 40-64 | 681 (33.5%) | 1.02 (0.8–1.2) | 0.823 | 681 (33.5%) | **1.3 (1.01–1.7)** | 0.040 |
|  | 65+ | 254 (12.5%) | 0.8 (0.6–1.1) | 0.227 | 254 (12.5%) | 1.4 (0.9–2.1) | 0.159 |
|  | Missing | 0 (0%) |  |  | 0 (0%) |  |  |
| Sex | Man | 458 (22.6%) | 1.0 (ref) |  | 458 (22.6%) | 1.0 (ref) |  |
|  | Female | 1573 (77.4%) | **1.3 (1.1–1.6)** | 0.007 | 1573 (77.4%) | **1.3 (1.1–1.7)** | 0.013 |
|  | Missing | 0 (0%) |  |  | 0 (0%) |  |  |
| Belonging to a vulnerable group | Yes | 1478 (72.8%) | 0.96 (0.8–1.2) | 0.658 | 1478 (72.8%) | 0.9 (0.8–1.2) | 0.559 |
|  | No | 553 (27.2%) | 1.0 (ref) |  | 553 (27.2%) | 1.0 (ref) |  |
|  | Missing | 0 (0%) |  |  | 0 (0%) |  |  |
| Education level | ≤ 11 years of schooling | 268 (13.3%) | 0.9 (0.7–1.2) | 0.476 | 268 (13.3%) | 1.04 (0.8–1.4) | 0.780 |
|  | Technical College or Vocational school | 554 (27.3%) | 0.9 (0.7–1.1) | 0.330 | 554 (27.3%) | 0.98 (0.8–1.2) | 0.828 |
|  | University degree and postgraduate degree | 1205 (59.2%) | 1.0 (ref) |  | 1205 (59.2%) | 1.0 (ref) |  |
|  | Missing | 4 (0.2%) |  |  | 4 (0.2%) |  |  |
| Settlement size | Regional center | 1392 (68.5%) | 1.0 (ref) |  | 1392 (68.5%) | 1.0 (ref) |  |
|  | City with more than 100 thousand inhabitants | 325 (16%) | 0.8 (0.7–1.1) | 0.184 | 325 (16%) | 0.98 (0.7–1.3) | 0.850 |
|  |  | N (%) | OR (95% CIs) | p-value | N (%) | AOR (95% CIs) | p-value |
|  | Settlement with less than 100 thousand inhabitants | 314 (15.5%) | 1.2 (0.9–1.5) | 0.152 | 314 (15.5%) | 1.2 (0.95–1.6) | 0.112 |
|  | Missing | 0 (0%) |  |  | 0 (0%) |  |  |
| **Number People in household** | 1-2 people | 611 (30.1%) | 1.0 (ref) |  | 611 (30.1%) | 1.0 (ref) |  |
|  | 3-4 people | 1199 (59%) | **1.4 (1.1–1.7)** | 0.002 | 1199 (59%) | **1.4 (1.1–1.8)** | 0.011 |
|  | 5+ people | 221 (10.9%) | 1.1 (0.8–1.5) | 0.482 | 221 (10.9%) | 1.01 (0.7–1.4) | 0.956 |
|  | Missing | 0 (0%) |  |  | 0 (0%) |  |  |
| **Income Level of the Household** | Can afford only the most necessary products | 773 (38.1%) | 0.9 (0.7–1.1) | 0.267 | 773 (38.1%) | 0.9 (0.7–1.2) | 0.432 |
|  | Can afford small additional items | 857 (42.2%) | 0.97 (0.8–1.2) | 0.796 | 857 (42.2%) | 1.01 (0.8–1.3) | 0.912 |
|  | Can afford everything | 401 (19.7%) | 1.0 (ref) |  | 401 (19.7%) | 1.0 (ref) |  |
|  | Missing | 0 (0%) |  |  | 0 (0%) |  |  |
| **Changes of Residency due to the Full-scale war** | Yes | 261 (12.8%) | 0.99 (0.8–1.3) | 0.963 | 261 (12.8%) | 1.1 (0.8–1.5) | 0.507 |
|  | No | 1766 (87%) | 1.0 (ref) |  | 1766 (87%) | 1.0 (ref) |  |
|  | Missing | 4 (0.2%) |  |  | 4 (0.2%) |  |  |
| **Employment status: Full-time work** | No | 1283 (63.2%) | 1.0 (ref) |  | 1283 (63.2%) | 1.0 (ref) |  |
|  | Yes | 748 (36.8%) | 0.9 (0.8–1.1) | 0.543 | 748 (36.8%) | 0.8 (0.6–1.01) | 0.063 |
|  | Missing | 0 (0%) |  |  | 0 (0%) |  |  |
|  | No | 1908 (93.9%) | 1.0 (ref) |  | 1908 (93.9%) | 1.0 (ref) |  |
|  |  | N (%) | OR (95% CIs) | p-value | N (%) | AOR (95% CIs) | p-value |
| **Employment status: Part-time work** | Yes | 123 (6.1%) | 1.1 (0.7–1.5) | 0.704 | 123 (6.1%) | 0.99 (0.7–1.5) | 0.952 |
|  | Missing | 0 (0%) |  |  | 0 (0%) |  |  |
| **Employment status: Self-employed** | No | 1953 (96.2%) | 1.0 (ref) |  | 1953 (96.2%) | 1.0 (ref) |  |
|  | Yes | 78 (3.8%) | 1.0 (0.6–1.6) | 0.996 | 78 (3.8%) | 0.8 (0.5–1.4) | 0.503 |
|  | Missing | 0 (0%) |  |  | 0 (0%) |  |  |
| **Employment status: Pensioner** | No | 1700 (83.7%) | 1.0 (ref) |  | 1700 (83.7%) | 1.0 (ref) |  |
|  | Yes | 331 (16.3%) | 0.8 (0.6–1.0) | 0.050 | 331 (16.3%) | 0.7 (0.5–1.05) | 0.083 |
|  | Missing | 0 (0%) |  |  | 0 (0%) |  |  |
| **Macro-region** | North | 336 (16.5%) | 1.3 (0.97–1.7) | 0.084 | 336 (16.5%) | 1.2 (0.9–1.6) | 0.191 |
|  | South | 239 (11.8%) | **1.6 (1.1–2.1)** | 0.005 | 239 (11.8%) | **1.6 (1.1–2.2)** | 0.009 |
|  | West | 517 (25.5%) | **2.0 (1.6–2.6)** | 0.000 | 517 (25.5%) | **2.1 (1.6–2.8)** | 0.000 |
|  | East | 476 (23.4%) | **2.0 (1.6–2.6)** | 0.000 | 476 (23.4%) | **1.9 (1.5–2.5)** | 0.000 |
|  | Center | 461 (22.7%) | 1.0 (ref) |  | 461 (22.7%) | 1.0 (ref) |  |
|  | Missing | 2 (0.1%) |  |  | 2 (0.1%) |  |  |
| **Changes in Access to Healthcare after Full-Scale War (24.02.2022)** | Yes, definitely in positive way | 216 (10.6%) | 1.0 (ref) |  | 216 (10.6%) | 1.0 (ref) |  |
|  | Yes, probably in positive way | 632 (31.2%) | 0.97 (0.7–1.3) | 0.865 | 632 (31.2%) | 0.7 (0.5–1.1) | 0.116 |
|  |  | N (%) | OR (95% CIs) | p-value | N (%) | AOR (95% CIs) | p-value |
|  | No changes | 809 (39.8%) | **1.6 (1.2–2.1)** | 0.004 | 809 (39.8%) | 0.9 (0.6–1.3) | 0.633 |
|  | No, probably in negative way | 145 (7.1%) | **1.9 (1.2–2.8)** | 0.004 | 145 (7.1%) | 0.9 (0.6–1.6) | 0.968 |
|  | No, definitely in negative way | 79 (3.9%) | **2.1 (1.2–3.5)** | 0.006 | 79 (3.9%) | 0.9 (0.5–1.7) | 0.740 |
|  | Difficult to say | 150 (7.4%) | 1.1 (0.7–1.7) | 0.694 | 150 (7.4%) | 0.7 (0.4–1.2) | 0.201 |
|  | Missing | 0 (0%) |  |  | 0 (0%) |  |  |
| **Changes in IPs in Healthcare after Full-Scale War (24.02.2022)** | Yes, definitely in positive way | 201 (9.9%) | 1.0 (ref) |  | 201 (9.9%) | 1.0 (ref) |  |
|  | Yes, probably in positive way | 601 (29.6%) | **1.5 (1.1–2.1)** | 0.013 | 601 (29.6%) | 1.4 (0.98–2.1) | 0.062 |
|  | No changes | 774 (38.1%) | **2.4 (1.7–3.3)** | 0.000 | 774 (38.1%) | **1.7 (1.2–2.6)** | 0.007 |
|  | No, probably in negative way | 110 (5.4%) | **3.2 (2.0–5.3)** | 0.000 | 110 (5.4%) | **2.5 (1.5–4.4)** | 0.001 |
|  | No, definitely in negative way | 109 (5.4%) | **3.4 (2.1–5.6)** | 0.000 | 109 (5.4%) | **2.3 (1.3–4.2)** | 0.005 |
|  | Difficult to say | 236 (11.6%) | **1.7 (1.1–2.4)** | 0.011 | 236 (11.6%) | 1.5 (0.95–2.5) | 0.078 |
|  | Missing | 0 (0%) |  |  | 0 (0%) |  |  |
| **Perceived success of healthcare reform in eliminating IPs** | Definitely possible | 205 (10.1%) | 1.0 (ref) |  | 205 (10.1%) | 1.0 (ref) |  |
|  | Probably possible | 747 (36.8%) | **1.5 (1.1–2.0)** | 0.020 | 747 (36.8%) | 1.4 (0.98–2.0) | 0.064 |
|  | Maybe possible, maybe no | 502 (24.7%) | **2.1 (1.5–2.9)** | 0.000 | 502 (24.7%) | **1.9 (1.3–2.7)** | 0.001 |
|  | Probably impossible | 212 (10.4%) | **2.6 (1.7–3.8)** | 0.000 | 212 (10.4%) | **2.3 (1.5–3.5)** | 0.000 |
|  | Definitely impossible | 249 (12.3%) | **3.7 (2.5–5.4)** | 0.000 | 249 (12.3%) | **3.0 (1.9–4.7)** | 0.000 |
|  |  | N (%) | OR (95% CIs) | p-value | N (%) | AOR (95% CIs) | p-value |
|  | Difficult to say | 116 (5.7%) | **1.7 (1.1–2.7)** | 0.029 | 116 (5.7%) | 1.6 (0.98–2.7) | 0.061 |

In the multivariable model, the following variables were significant: age group 40-64 years old (AOR 1.3, 95% CI 1.01-1.7); female participants (AOR 1.3, 95% CI 1.1-1.7); households with 3-4 people (AOR 1.4, 95% CI 1.1-1.8); lived in southern (AOR 1.6, 95% CI 1.1-2.1), western (AOR 2.1, 95% CI 1.6-2.8), eastern (AOR 1.9, 95% CI 1.5-2.5) macro-regions of Ukraine; faced changes in IPs after full-scale war started in probably negative way (OR 2.5, 95% CI 1.5-4.4), in definitely negative way (AOR 2.3, 95% CI 1.3-4.2), no changes (AOR 1.7, 95% CI 1.2-2.6); evaluated expected overcoming of IPs in healthcare as probably impossible (AOR 2.3, 95% CI 1.5-3.5), definitely impossible (AOR 3.0, 95% CI 2.0-4.7), maybe possible, maybe no (AOR 1.9, 95% CI 1.3-2.7).

### Informal payments within last 12 months: Did you make an informal payment for primary care last year?

Overall, 45.5% (95% CIs: 43.3-47.7) of patients made the IPs at the PHC level last year. In the bivariate model (Table 3), the following variables were associated with attitude towards IPs: female participants (OR 1.4, 95% CI 1.1-1.7); households with 3-4 people (OR 1.4, 95% CI 1.2-1.8); lived in southern (OR 1.5, 95% CI 1.1-2.1), western (OR 2.0, 95% CI 1.6-2.6), eastern (OR 1.9, 95% CI 1.5-2.5) macro-regions of Ukraine; experienced changes in IPs since full-scale war in a probably positive way (OR 1.5, 95% CI 1.1-2.1), probably negative way (OR 3.2, 95% CI 2.0-5.2), definitely negative way (OR 3.6, 95% CI 2.2-5.8), no changes (OR 2.4, 95% CI 1.7-3.3), or uncertain (OR 1.6, 95% CI 1.1-2.5); evaluated expected overcoming of IPs in healthcare as probably possible (OR 1.5, 95% CI 1.05-2.0), probably impossible (OR 2.8, 95% CI 1.9-4.2), definitely impossible (OR 4.0, 95% CI 2.7-5.9), maybe possible, maybe no (OR 2.2, 95% CI 1.5-3.0), difficult to say (OR 1.8, 95% CI 1.1-2.8).

**Table 3.** Bivariate and Multivariable Analysis of Informal Payments for Primary Health Care Services with the Last 12 Months.

| Independent variables |  | Bivariate analysis |  |  | Multivariable analysis |  |  |
| --- | --- | --- | --- | --- | --- | --- | --- |
|  |  | N (%) | OR (95% CIs) | p-value | N (%) | AOR (95% CIs) | p-value |
| **Age in years** | 18-39 | 1096 (54%) | 1.0 (ref) |  | 1096 (54%) | 1.0 (ref) |  |
|  | 40-64 | 681 (33.5%) | 0.99 (0.8–1.2) | 0.889 | 681 (33.5%) | 1.2 (0.96–1.6) | 0.097 |
|  | 65+ | 254 (12.5%) | 0.8 (0.6–1.04) | 0.093 | 254 (12.5%) | 1.2 (0.8–1.9) | 0.380 |
|  | Missing | 0 (0%) |  |  | 0 (0%) |  |  |
| **Sex** | Man | 458 (22.6%) | 1.0 (ref) |  | 458 (22.6%) | 1.0 (ref) |  |
|  | Female | 1573 (77.4%) | **1.4 (1.1–1.7)** | 0.003 | 1573 (77.4%) | **1.4 (1.1–1.8)** | 0.007 |
|  | Missing | 0 (0%) |  |  | 0 (0%) |  |  |
| **Belonging to a vulnerable group** | No | 1478 (72.8%) | 1.01 (0.8–1.2) | 0.961 | 1478 (72.8%) | 1.01 (0.8–1.3) | 0.948 |
|  | Yes | 553 (27.2%) | 1.0 (ref) |  | 553 (27.2%) | 1.0 (ref) |  |
|  | Missing | 0 (0%) |  |  | 0 (0%) |  |  |
| **Education level** | ≤ 11 years of schooling | 268 (13.3%) | 0.9 (0.7–1.2) | 0.541 | 268 (13.3%) | 1.1 (0.8–1.4) | 0.628 |
|  | Technical College or Vocational school | 554 (27.3%) | 0.9 (0.7–1.1) | 0.329 | 554 (27.3%) | 0.99 (0.8–1.2) | 0.984 |
|  | University degree and postgraduate degree | 1205 (59.2%) | 1.0 (ref) |  | 1205 (59.2%) | 1.0 (ref) |  |
|  | Missing | 4 (0.2%) |  |  | 4 (0.2%) |  |  |
| **Settlement size** | Regional center | 1392 (68.5%) | 1.0 (ref) |  | 1392 (68.5%) | 1.0 (ref) |  |
|  | City with more than 100 thousand inhabitants | 325 (16%) | 0.8 (0.6–1.03) | 0.087 | 325 (16%) | 0.9 (0.7–1.2) | 0.462 |
|  |  | N (%) | OR (95% CIs) | p-value | N (%) | AOR (95% CIs) | p-value |
|  | Settlement with less than 100 thousand inhabitants | 314 (15.5%) | 1.2 (0.9–1.5) | 0.226 | 314 (15.5%) | 1.2 (0.9–1.6) | 0.190 |
|  | Missing | 0 (0%) |  |  | 0 (0%) |  |  |
| **Number People in household** | 1-2 people | 611 (30.1%) | 1.0 (ref) |  | 611 (30.1%) | 1.0 (ref) |  |
|  | 3-4 people | 1199 (59%) | **1.4 (1.2–1.8)** | 0.000 | 1199 (59%) | **1.5 (1.1–1.9)** | 0.005 |
|  | 5+ people | 221 (10.9%) | 1.2 (0.9–1.7) | 0.231 | 221 (10.9%) | 1.1 (0.8–1.5) | 0.679 |
|  | Missing | 0 (0%) |  |  | 0 (0%) |  |  |
| **Income Level of the Household** | Can afford only the most necessary products | 773 (38.1%) | 0.9 (0.7–1.1) | 0.215 | 773 (38.1%) | 0.9 (0.7–1.2) | 0.362 |
|  | Can afford small additional items | 857 (42.2%) | 0.96 (0.8–1.2) | 0.724 | 857 (42.2%) | 1.0 (0.8–1.3) | 0.998 |
|  | Can afford everything | 401 (19.7%) | 1.0 (ref) |  | 401 (19.7%) | 1.0 (ref) |  |
|  | Missing | 0 (0%) |  |  | 0 (0%) |  |  |
| **Changes of Residency due to the Full-scale war** | Yes | 261 (12.8%) | 0.96 (0.7–1.3) | 0.758 | 261 (12.8%) | 1.1 (0.8–1.4) | 0.653 |
|  | No | 1766 (87%) | 1.0 (ref) |  | 1766 (87%) | 1.0 (ref) |  |
|  | Missing | 4 (0.2%) |  |  | 4 (0.2%) |  |  |
| **Employment status: Full-time work** | No | 1283 (63.2%) | 1.0 (ref) |  | 1283 (63.2%) | 1.0 (ref) |  |
|  | Yes | 748 (36.8%) | 0.96 (0.8–1.1) | 0.636 | 748 (36.8%) | 0.8 (0.7–1.05) | 0.118 |
|  | Missing | 0 (0%) |  |  | 0 (0%) |  |  |
|  |  | N (%) | OR (95% CIs) | p-value | N (%) | AOR (95% CIs) | p-value |
| **Employment status: Part-time work** | No | 1908 (93.9%) | 1.0 (ref) |  | 1908 (93.9%) | 1.0 (ref) |  |
|  | Yes | 123 (6.1%) | 1.02 (0.7–1.5) | 0.935 | 123 (6.1%) | 0.9 (0.6–1.4) | 0.805 |
|  | Missing | 0 (0%) |  |  | 0 (0%) |  |  |
| **Employment status: Self-employed** | No | 1953 (96.2%) | 1.0 (ref) |  | 1953 (96.2%) | 1.0 (ref) |  |
|  | Yes | 78 (3.8%) | 0.98 (0.6–1.6) | 0.946 | 78 (3.8%) | 0.9 (0.5–1.4) | 0.551 |
|  | Missing | 0 (0%) |  |  | 0 (0%) |  |  |
| **Employment status: Pensioner** | No | 1700 (83.7%) | 1.0 (ref) |  | 1700 (83.7%) | 1.0 (ref) |  |
|  | Yes | 331 (16.3%) | 0.8 (0.6–0.98) | 0.031 | 331 (16.3%) | 0.7 (0.5–1.1) | 0.143 |
|  | Missing | 0 (0%) |  |  | 0 (0%) |  |  |
| **Macro region** | North | 336 (16.5%) | 1.2 (0.9–1.6) | 0.294 | 336 (16.5%) | 1.1 (0.8–1.5) | 0.547 |
|  | South | 239 (11.8%) | **1.5 (1.1–2.1)** | 0.012 | 239 (11.8%) | **1.5 (1.1–2.2)** | 0.017 |
|  | West | 517 (25.5%) | **2.0 (1.6–2.6)** | 0.000 | 517 (25.5%) | **2.1 (1.6–2.8)** | 0.000 |
|  | East | 476 (23.4%) | **1.9 (1.5–2.5)** | 0.000 | 476 (23.4%) | **1.8 (1.4–2.4)** | 0.000 |
|  | Center | 461 (22.7%) | 1.0 (ref) |  | 461 (22.7%) | 1.0 (ref) |  |
|  | Missing | 2 (0.1%) |  |  | 2 (0.1%) |  |  |
|  | Yes, definitely in positive way | 216 (10.6%) | 1.0 (ref) |  | 216 (10.6%) | 1.0 (ref) |  |
|  | Yes, probably in positive way | 632 (31.2%) | 1.03 (0.7–1.4) | 0.856 | 632 (31.2%) | 0.8 (0.6–1.2) | 0.338 |
|  |  | N (%) | OR (95% CIs) | p-value | N (%) | AOR (95% CIs) | p-value |
| **Changes in Access to Healthcare after Full-Scale War (24.02.2022)** | No changes | 809 (39.8%) | 1.7 (1.2-2.3) | 0.001 | 809 (39.8%) | 1.01 (0.7–1.5) | 0.975 |
|  | No, probably in negative way | 145 (7.1%) | 2.0 (1.3-3.1) | 0.001 | 145 (7.1%) | 1.1 (0.7–1.9) | 0.686 |
|  | No, definitely in negative way | 79 (3.9%) | 2.3 (1.3-3.9) | 0.002 | 79 (3.9%) | 0.96 (0.5–1.9) | 0.910 |
|  | Difficult to say | 150 (7.4%) | 1.2 (0.8-1.9) | 0.376 | 150 (7.4%) | 0.8 (0.5–1.4) | 0.454 |
|  | Missing | 0 (0%) |  |  | 0 (0%) |  |  |
| **Changes in IPs in Healthcare after Full-Scale War (24.02.2022)** | Yes, definitely in positive way | 201 (9.9%) | 1.0 (ref) |  | 201 (9.9%) | 1.0 (ref) |  |
|  | Yes, probably in positive way | 601 (29.6%) | **1.5 (1.04–2.1)** | 0.029 | 601 (29.6%) | 1.3 (0.9–1.9) | 0.194 |
|  | No changes | 774 (38.1%) | **2.4 (1.7–3.3)** | 0.000 | 774 (38.1%) | **1.6 (1.1–2.4)** | 0.023 |
|  | No, probably in negative way | 110 (5.4%) | **3.2 (2.0–5.2)** | 0.000 | 110 (5.4%) | **2.4 (1.4–4.2)** | 0.003 |
|  | No, definitely in negative way | 109 (5.4%) | **3.6 (2.2–5.8)** | 0.000 | 109 (5.4%) | **2.2 (1.2–4.1)** | 0.008 |
|  | Difficult to say | 236 (11.6%) | **1.6 (1.1–2.5)** | 0.014 | 236 (11.6%) | 1.4 (0.9–2.3) | 0.168 |
|  | Missing | 0 (0%) |  |  | 0 (0%) |  |  |
| **Perceived success of healthcare reform in eliminating IPs** | Definitely possible | 205 (10.1%) | 1.0 (ref) |  | 205 (10.1%) | 1.0 (ref) |  |
|  | Probably possible | 747 (36.8%) | **1.5 (1.05–2.0)** | 0.025 | 747 (36.8%) | 1.4 (0.99–2.0) | 0.060 |
|  | Maybe possible, maybe no | 502 (24.7%) | **2.2 (1.5–3.0)** | 0.000 | 502 (24.7%) | **2.0 (1.3–2.9)** | 0.001 |
|  | Probably impossible | 212 (10.4%) | **2.8 (1.9–4.2)** | 0.000 | 212 (10.4%) | **2.5 (1.6–3.9)** | 0.000 |
|  | Definitely impossible | 249 (12.3%) | **4.0 (2.7–5.9)** | 0.000 | 249 (12.3%) | **3.4 (2.1–5.2)** | 0.000 |
|  |  | N (%) | OR (95% CIs) | p-value | N (%) | AOR (95% CIs) | p-value |
|  | Difficult to say | 116 (5.7%) | **1.8 (1.1–2.8)** | 0.020 | 116 (5.7%) | **1.7 (1.04–2.9)** | 0.036 |
|  | Missing | 0 (0%) |  |  | 0 (0%) |  |  |

In the final multivariable model, the following variables were independently associated with: female participants (AOR 1.4, 95% CI 1.1-1.8); have 3-4 people in the household (AOR 1.5, 95% CI 1.1-1.9); lived in southern (AOR 1.5, 95% CI 1.1-2.2), western (AOR 2.1, 95% CI 1.6-2.8), eastern (AOR 1.8, 95% CI 1.4-2.4) macro-regions of Ukraine; faced changes in IPs since the full-scale war started in probably negative way (AOR 2.4, 95% CI 1.4-4.2), in a definitely negative way (AOR 2.2, 95% CI 1.2-4.1), no changes (AOR 1.6, 95% CI 1.1-2.4); evaluated expected overcoming of IPs in healthcare as probably impossible (AOR 2.5, 95% CI 1.6-3.9), definitely impossible (AOR 3.4, 95% CI 2.1-5.2), maybe possible, maybe no (AOR 2.5, 95% CI 1.6-3.9), difficult to say (AOR 1.7, 95% CI 1.04-2.9).

### Attitude to informal payments in healthcare

In total, 62.5% (95% CIs: 60.3-64.6) of participants reported a negative attitude toward IPs in healthcare (Table 4). In the bivariate model, a negative attitude toward IPs was more likely among individuals aged 40-64 years (OR 0.5, 95% CI 0.4-0.7) and those aged 65+ years (OR 0.4, 95% CI 0.3-0.5). It was also common among residents of smaller settlements (OR 0.6, 95% CI 0.5-0.8), people who can afford only basic necessities (OR 0.5, 95% CI 0.4-0.6) or small extras (OR 0.7, 95% CI 0.6-0.9), non-working pensioners (OR 0.4, 95% CI 0.3-0.6), residents of northern macro-region of Ukraine (OR 0.7, 95% CI 0.5-0.96), and those who recognized negative changes in healthcare access after the full-scale war began (OR 0.5, 95% CI 0.2-0.8). On the other hand, inhabitants of cities with populations over 100,000 (OR 1.5, 95% CI 1.2-1.9), households with 3-4 members (OR 1.6, 95% CI 1.3-2.0), and households with 5 or more members (OR 1.8, 95% CI 1.3-2.5) were more likely to have a positive attitude toward IPs.

**Table 4.**
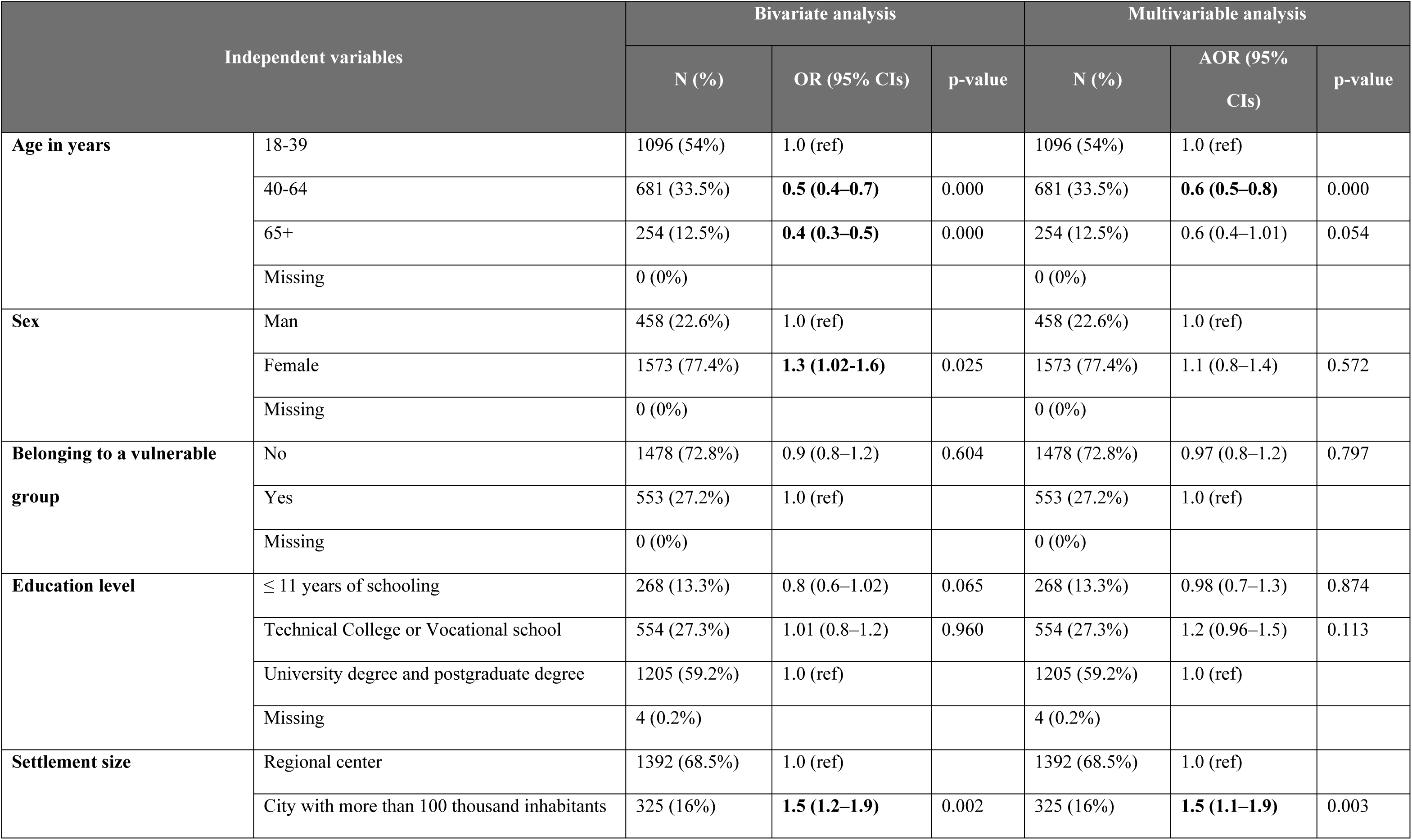
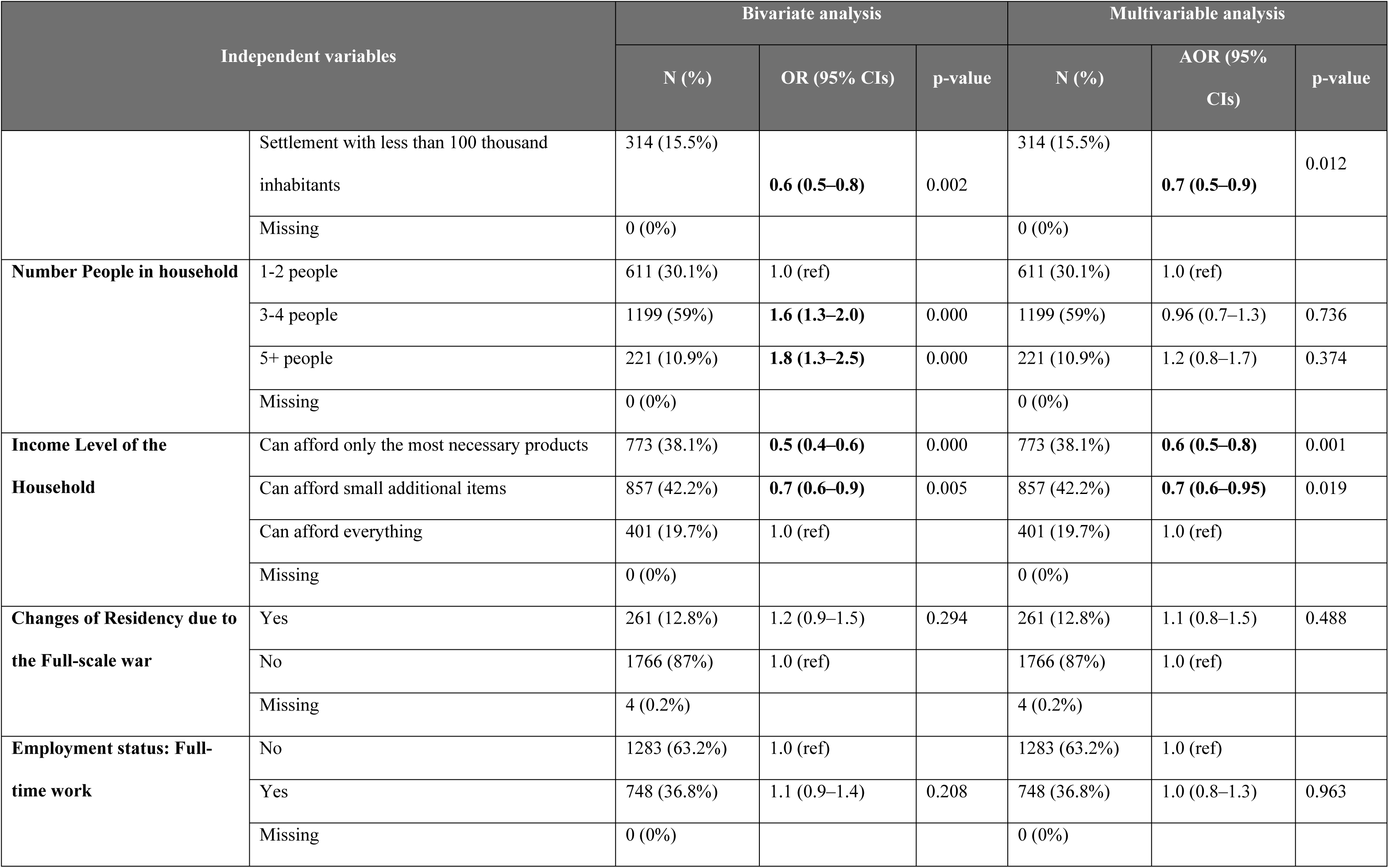
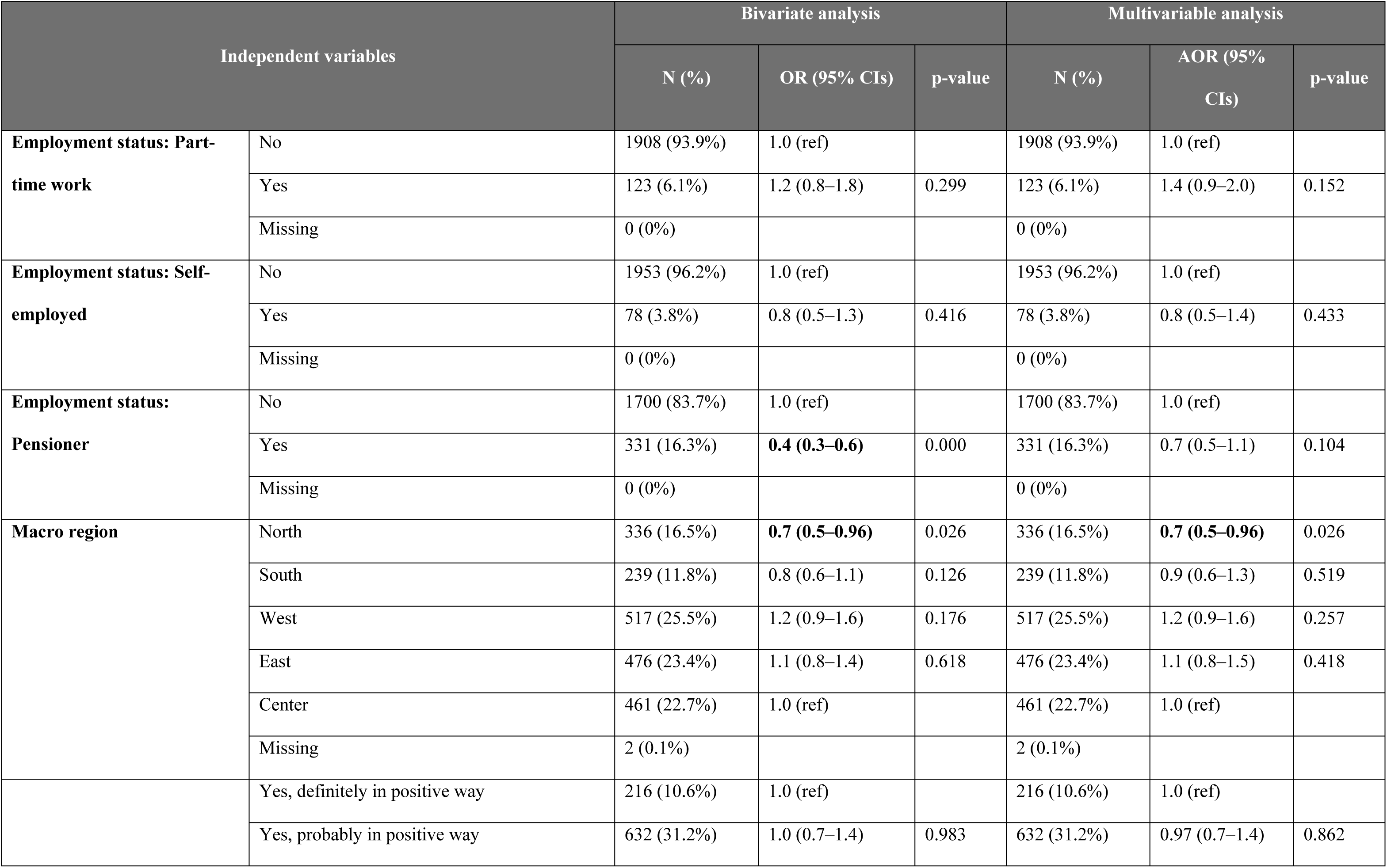
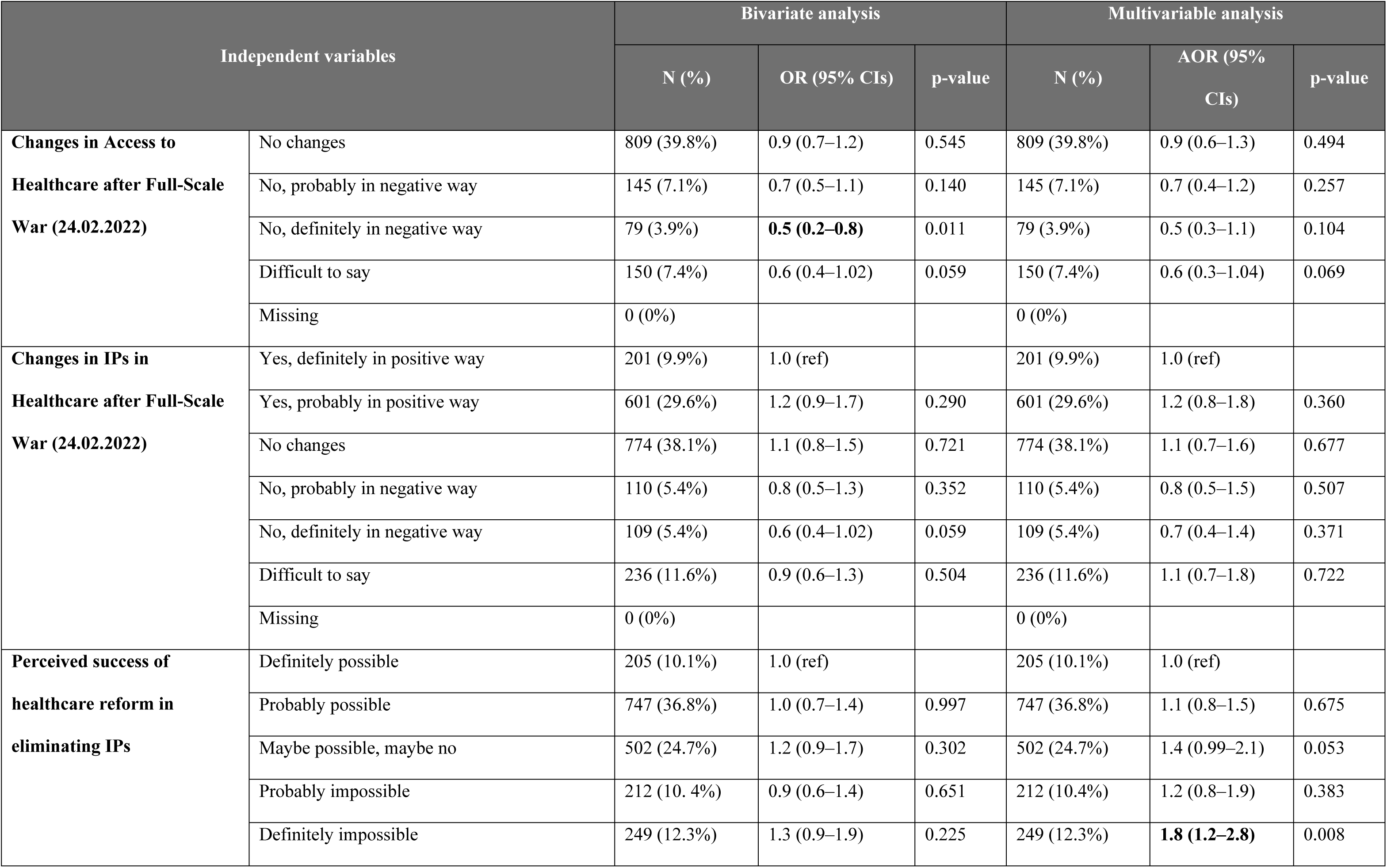
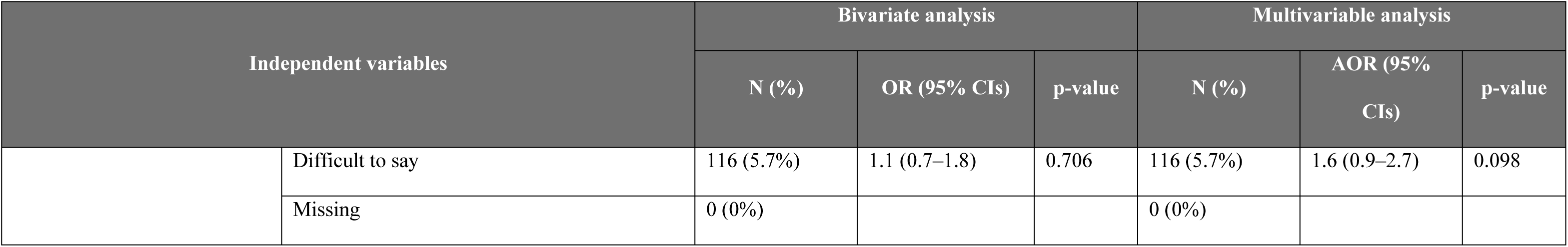
Bivariate and Multivariable Analysis of Patient Attitudes Toward Informal Payments in Ukraine.

In the multivariable model, a negative association with IPs was observed among residents of smaller settlements (AOR 0.7, 95% CI 0.5-0.9), those who can afford only basic necessities (AOR 0.6, 95% CI 0.5-0.8), individuals aged 40-64 years (AOR 0.6, 95% CI 0.5-0.8), those who can afford small extras (AOR 0.7, 95% CI 0.6-0.9), and residents of northern macro-region of Ukraine (AOR 0.7, 95% CI 0.5-0.96).

The following variables were associated with positive attitude towards IPs; inhabitants of cities with more than 100 000 people (AOR 1.5, 95% CI 1.1-1.9); thinking that it is definitely impossible to overcome IPs in healthcare (AOR 1.8, 95% CI 1.2-2.8). Participants were more likely to have a positive attitude toward IPs if they lived in cities with populations over 100,000 (AOR 1.5, 95% CI 1.1-1.9) or believed it was impossible to eliminate IPs in healthcare (AOR 1.8, 95% CI 1.2-2.8).

## Discussion

In this study, we analysed the factors associated with the lifetime occurrence of IPs on PHC level, the recent IPs within the past 12 months on PHC level, and prevailing attitudes toward this practice among patients receiving the four key healthcare services in Ukraine: childbirth, neonatal care, acute myocardial infarction, and acute stroke care. The findings provide critical insights into the factors that are linked to an increased likelihood of making IPs and the development of positive attitudes toward this practice.

Our study demonstrated that female patients were more likely to have made IPs both over their lifetime and within the past 12 months on PHC level. This finding aligns with previous research that suggested that women were more likely to engage in IPs, potentially due to their higher utilization of healthcare services. Thus, it has been shown that women tend to have more frequent doctor visits and a greater demand for diagnostic and therapeutic interventions, particularly in relation to reproductive health, leading to increased interactions with healthcare providers and, consequently, a higher likelihood of making IPs [37–41]. Additionally, sociocultural factors, including the desire to maintain positive relationships with healthcare professionals, may further contribute to this trend [11, 37, 39]. Women may feel particularly compelled to ensure high-quality care during critical periods such as pregnancy and childbirth, which can drive IPs aimed at securing expedited or enhanced services and their quality. This behaviour is especially prevalent in contexts where healthcare quality is perceived as inadequate without such payments [42]. In this context, it is important to note that evidence shows women in the United States make approximately 80% of healthcare decisions for their families, yet often lack adequate healthcare coverage themselves [43].Thus, it is crucial to prioritize women in the design of information campaigns, ensuring they have access to reliable, up-to-date information and support, particularly during critical life stages such as pregnancy and childbirth. Furthermore, future research should focus on systematically examining the gender-specific needs and vulnerabilities that women in Ukraine face with respect to IPs. Such insights are crucial for developing more precisely targeted interventions to effectively address and mitigate these challenges.

We also observed an association between the number of people in the household and the likelihood of IPs, which similarly might be explained by increased utilization of healthcare services and more frequent engagement with healthcare workers. To this end it is also crucial to mention that 80% of women are responsible for household healthcare decisions (Matoff-Stepp et al., 2014).

Previous research suggests that older patients tend to make fewer IPs [42]. Although we did not observe any significant association between age and the likelihood of making IPs either over a lifetime or in the past 12 months (potentially due to the way age groups were categorized) on PHC level, we found that older patients generally held negative attitudes towards IPs. This may be explained by the fact that older individuals tend to be more financially vulnerable and have greater exposure to the healthcare system, owing to higher utilization of services and longer periods of healthcare use throughout their lives compared to younger individuals [42]. It is important to examine whether, with age, patients in Ukraine tend to make informal payments less or whether their negative attitudes towards IPs fail to translate into changes in their behaviour. Additionally, the prevalent negative attitude among older patients could be leveraged in healthcare service monitoring, as these individuals may potentially serve as valuable sources of information regarding IP practices.

Moreover, our results demonstrated that individuals with lower income levels were more likely to hold negative attitudes toward IPs, which might be due to the catastrophic financial burden such payments impose on low-income households [14, 44, 45]. This challenge is further compounded by prior research showing that in countries like Ukraine and Romania, where IPs are prevalent and poverty rates are relatively high, patients bear a disproportionately heavy burden and difficulties in paying for healthcare services [14]. Therefore, households with low-income levels should be prioritized and targeted by state programs to ensure that healthcare services are inclusive, accessible to all, and aligned with the goal of achieving universal health coverage.

Also, our results suggested that individuals who did not support ongoing healthcare reforms, including those aimed at more efficient and transparent resource allocation, strategic purchasing with a purchaser-provider split, clearly defined health benefits, and new payment methods for healthcare providers – were more likely to have a positive attitude towards IPs and had higher odds of making informal payments for healthcare services both over their lifetime and within the past 12 months. So, individuals with negative attitudes toward healthcare reforms and ongoing changes may be less likely to alter their behaviour or beliefs regarding IPs. This finding can also be interpreted through the lens of prevailing social beliefs and norms regarding IPs and healthcare services in Ukraine and other Central and Eastern European countries, where such payments are often culturally ingrained [11–13]. Overall, studies from these regions have demonstrated that, despite a general desire to address the issue of IPs, the public frequently see these payments as necessary to obtain more personalized care, improved quality, and expedited access to healthcare services [14, 46–48]. We emphasize the need for a more nuanced understanding of how social norms and health beliefs shape patients’ attitudes and behaviours toward IPs. These factors must be carefully considered when developing healthcare service delivery strategies.

Although no associations were observed between outcome variables and changes in residency due to the full-scale war, patients who reported no changes in informal payments or a worsening situation after the war were more likely to have made payments in the past 12 months and at some point, in their lives on PHC level. This is most likely linked to the overarching health beliefs and behaviours of these respondents, but it could also be a direct effect of the war. Future research should carefully investigate how, and to what extent, the full-scale war has influenced the behaviour of both patients and providers regarding informal payments in Ukraine.

Finally, another interesting finding is the regional variation in the likelihood of IPs, particularly between the central macro-region with Kyiv city and other macro-regions (i.e., southern, northern, western, and eastern). This may be attributed to differences in the timing of healthcare reforms as well as the effectiveness of their implementation and later enforcement on the ground, leading to varying outcomes in these localities. Additionally, the number of people residing in rural areas may also influence these associations. There is evidence suggesting that individuals in rural areas may be more willing to make IPs [13, 49–52]. However, it is also important to consider that this willingness might be influenced by the low-income levels prevalent in the rural areas [13, 35, 50, 53]. We thus, argue that regional differences may play a crucial role in shaping context-specific and more efficient approaches to healthcare service delivery.

### Future studies

A third Informal Payment Study focused on the same priority health services should be conducted within three years. The study’s design and methodology should be adjusted to reflect conditions relevant to the new time period and new trends in healthcare system in Ukraine.

### Recommendations

To address the factors influencing IPs in Ukraine’s healthcare system, a range of targeted interventions and systemic improvements is essential:

- A phased approach should be used to incorporate study results into the calculation of tariffs for priority health services under the state-funded Program of Medical Guarantees, ensuring tariffs reflect current healthcare conditions.
- Public sector healthcare providers should focus on establishing competitive salaries for healthcare staff by improving financial resource efficiency, which may help reduce the demand for IPs.
- Service provision for vulnerable groups, particularly those at risk of catastrophic expenditures, should be considered while designing health programs at all levels – state, regional and local.
- Gender-sensitive strategies should prioritize women, particularly during critical life stages like pregnancy, providing clear information on healthcare entitlements to reduce reliance on IPs.
- Low-income households who bear a disproportionate financial burden from IPs, should receive targeted support through state assistance programs aligned with UHC goals.
- National and local authorities should lead public education campaigns to highlight the benefits of healthcare reforms, discourage IP practices, and shift social norms that perpetuate IPs. Health programs should ensure clear, detailed information on service delivery for each priority service.

## Conclusion

This study provides valuable insights into the prevalence and determinants of IPs within Ukraine’s primary healthcare system, focusing on the key health services such as childbirth, neonatal care, and acute care for myocardial infarction and stroke. The findings highlight critical factors associated with the likelihood of making IPs, including sex, income, household size, and attitudes toward healthcare reforms. The widespread and normalized use of IPs among both healthcare providers and patients over the years has led to a significant financial burden, particularly on individuals with lower incomes and greater healthcare needs.

One of the key policy responses to address IPs has been the inclusion of these four priority health services in the Program of Medical Guarantees that ensured that payments for these services are made directly to healthcare institutions by the NHSU. This policy is already being implemented and expands financial risk protection for vulnerable groups, including low-income populations, by reducing their reliance on informal payments.

The elimination of IPs remains a central objective of ongoing health reforms in Ukraine, a goal that is particularly important in the context of the current full-scale war. The anticipated impacts of eliminating IPs include removing financial barriers to care, promoting equitable access to healthcare services, reducing corruption in publicly funded facilities, and enhancing financial risk protection for patients. Indirectly, this policy may also contribute to poverty reduction by preventing catastrophic health expenditures, particularly among vulnerable groups who face both health and social disadvantages. Initial signs of progress in the reform suggest improvements in health financing, funding, and the development of integrated, transparent healthcare operations.

Complementary strategies to eradicate IPs should include improving the working conditions and salaries of healthcare staff, as well as implementing educational campaigns for both healthcare providers and patients to raise awareness about the illegality of IPs. The findings of this study are essential for informing evidence-based policy decisions and communication strategies designed to combat IPs and further Ukraine’s health reform objective of providing financial risk protection against catastrophic health expenditures.

Overall, the results of this study contribute to the formulation of data-driven interventions and social and behavioural change communication strategies. These efforts enable the monitoring of IP practices and support the achievement of health reform goals that seek to reduce corruption and improve health outcomes. In conclusion, addressing IPs within Ukraine’s healthcare system requires a comprehensive, context-specific approach, which includes targeted reforms, effective public awareness campaigns, and strategic resource allocation. By addressing the root causes of IPs and implementing evidence-based policy changes, Ukraine can progress toward a more transparent and equitable healthcare system, reducing financial barriers to care and ensuring universal access to essential services.

## Statements and Declarations

## Author Contributions

“All authors contributed to the study’s conception and design. Maksym Duda and Nataliia Kovalenko led the survey design and supervision. Material preparation and data analysis were conducted by Maksym Duda, Nataliia Kovalenko, Anna Tokar, Oleh Sorotsynskyi, Paola Pavlenko and Vladyslav Odrynskyi. Maksym Duda, Anna Tokar, and Oleh Sorotsynskyi wrote the first draft of the manuscript. Anna Tokar and Oleh Sorotsynskyi performed the statistical analysis. All authors provided feedback on previous versions of the manuscript and approved the final version.”

## Data Availability

The data underlying the results presented in the study are available from the Kyiv School of Economics (KSE), Ukraine and the first author - Dr Maksym Duda.

## Acknowledgement

We would like to acknowledge the Kyiv School of Economics (KSE) for leading the primary data collection and analysis conducted as part of the USAID Health Reform Support project.

## Statements and Declarations

